# Medication Exposure and Neurodegenerative Disease Risk Across National Biobanks

**DOI:** 10.1101/2025.07.07.25330740

**Authors:** Kristin S. Levine, Lana Sargent, Emma N. Somerville, Vanessa Pitz, Elvin T. Price, Sara Bandres-Ciga, Emily Simmonds, Rodney Alan Long, Caroline Jonson, Lara M. Lange, Alastair J Noyce, Valentina Escott-Price, Hirotaka Iwaki, Kendall Van Keuren-Jensen, Luigi Ferrucci, Mark R Cookson, Andrew Singleton, Mike Nalls, Hampton Leonard

## Abstract

**Abstract:** Advanced age, genetics, and environmental exposures are leading contributors to the development of neurodegenerative disorders (NDD). In this study, we used data from the UK Biobank (UKB) and the All of Us (AoU) initiative to determine if exposure to specific medications are associated with an increased or decreased risk of NDD, including Alzheimer’s (AD), Parkinson’s disease (PD), and all-cause dementia (DEM). We investigated the associations between these diseases and prescription drug exposures through an unbiased analysis, assessing both lifetime risk and risk from exposures occurring more than ten years before diagnosis, while also accounting for comorbid conditions.

**Methods:** Cox proportional hazard models were used to evaluate both lifetime and ten-year lag-associated risks of developing a NDD following exposure to specific prescription medications. This analysis followed a two-stage design, incorporating separate discovery and replication cohorts sourced from national-scale biobanks.

**Findings:** We pulled data from over 700,000 health records from individuals of European ancestry to survey a total of 480 prescription medication exposures. After multiple test corrections, we found 241 significant associations between medication exposure and risk of NDDs in our discovery cohort, with 157 of these replicated in an independent dataset. After adjusting for potential comorbidities, 15 medication-NDD associations remained significant, some of which were attenuated after accounting for *APOE*-ε4 status. Most of these significant pairings were associated with increased risk, however, two antibiotics, one proton pump inhibitor, and one statin had replicated effects inversely associated with disease risk.

**Interpretation:** While correlation does not imply causation and some associations may be driven by medications used to treat prodromal stages of disease, we have utilized large, unbiased datasets to identify and replicate associations between commonly prescribed medications and NDD risk. Additional longitudinal and mechanistic investigations are warranted.

## Introduction

Older adults diagnosed with a neurodegenerative disease (NDD) often have other comorbid diseases such as diabetes, cardiovascular diseases, depression, or anxiety.^1,2^ Concomitant diseases often necessitate the prescription of multiple medications leading to potential medication interactions. Several medications used in NDDs, including antidepressants, antipsychotics, and benzodiazepines, are known to have adverse clinical reactions, including effects on cognitive function. ^3,4,5^

Previous research has identified a number of medication exposures that may impose risk or protection for the later development of neurodegenerative diseases, with studies finding contradictory results. ^6,7^ The use of statins and the risk of Alzheimer’s Disease (AD) is one example of this, with multiple observational studies and systematic reviews reporting protective effects of statins,^8–10^ but clinical trials have so far failed to recreate this effect in AD patients, although there have been some positive trial findings for mild cognitive impairment (MCI).^11,12^ With the recent availability of biobank-scale data, we sought to examine potential associations between medication exposure(s) and NDD risk across large populations at different time periods and different prescription quantities, aiming to provide additional context for long-term medication use.

In this study, we used data from the UK Biobank (UKB) and the All of Us (AoU) initiative to identify potential associations between medication exposures and a variety of common NDDs including AD, all-cause dementia (DEM), and Parkinson’s disease (PD). This report surveyed longitudinal and cross-sectional associations between medication exposures and NDDs in a hypothesis-free assessment, looking at all medication data available across the UKB and AoU. In addition, we investigated associations with additional models using comorbid disease diagnosis (ICD10 codes) and *APOE*-ε4 status, testing for confounding factors for some of these medication exposure associations. A summary of the study design, sample sizes with available medication data (see methods section for exclusion criteria), and analyses can be found in **Figure 1**.

**Figure 1:**
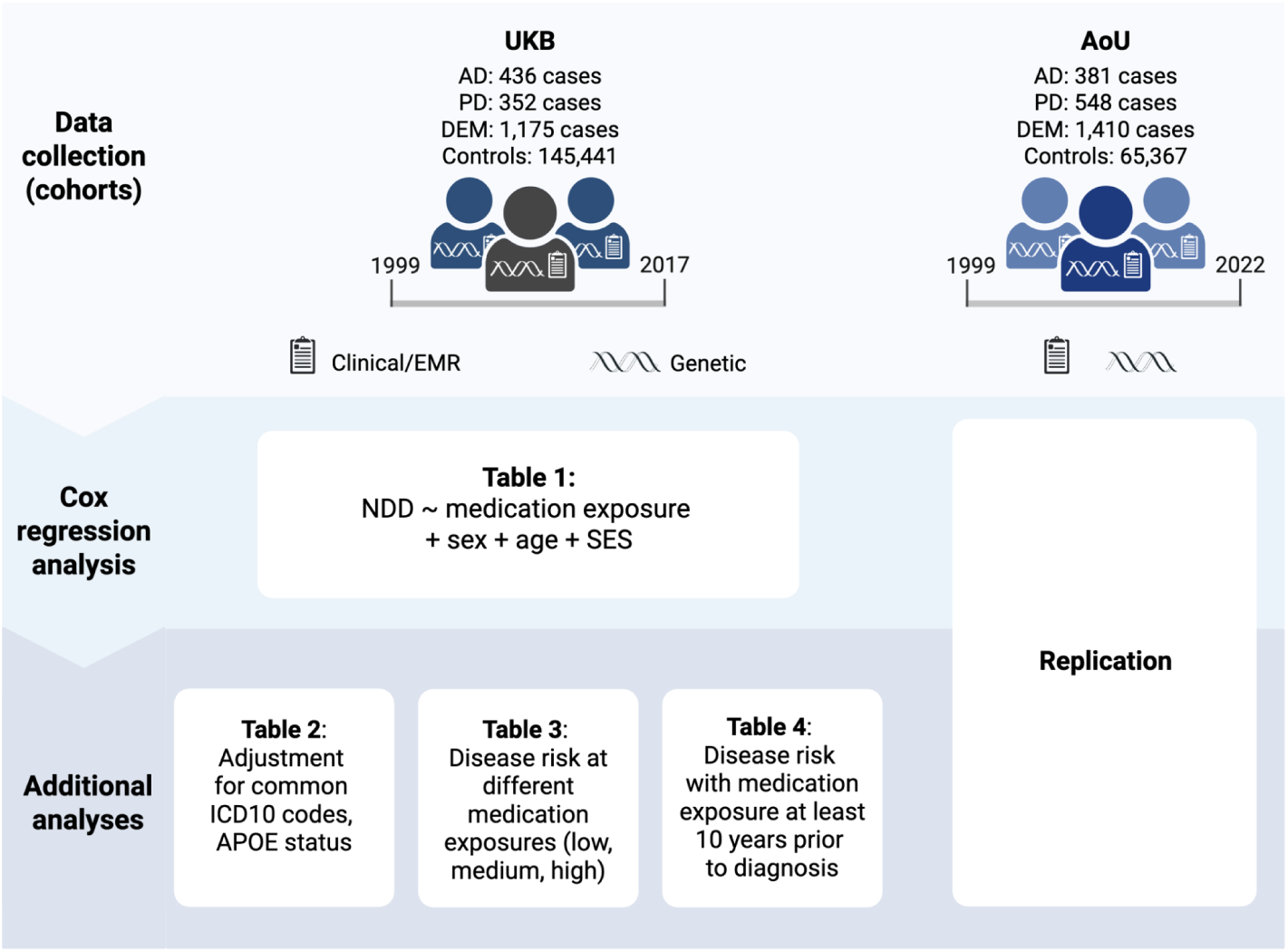
Summary of Study Design and Analyses.

## Results

### Associations between any medication exposure and neurodegeneration

Using biobank-scale data, we employed a two-stage study design to investigate associations between common medication use and the risk of incident cases of AD, DEM, and PD. The discovery phase was conducted in UKB, followed by replication in AoU. We tested 480 medication–disease pairings in UKB, of which 241 passed false discovery rate (FDR) correction (detailed in <u>Table S1</u>). All analyses were right-censored to include only medications first taken prior to NDD diagnosis. Of the 241 significant associations, 157 replicated in AoU with hazard ratios (HRs) showing consistent directionality (<u>Table S1</u>). A total of 96 unique medications were associated with one or more diseases of interest (<u>Table S1</u>).

Of the 96 unique medications with replicated associations, 47 were associated with two or more NDDs; 14 were associated with all three NDDs (total significant associations across the two-stages of analysis, N = 157). See **Figure 2** for a Venn Diagram of these results.

**Figure 2:**
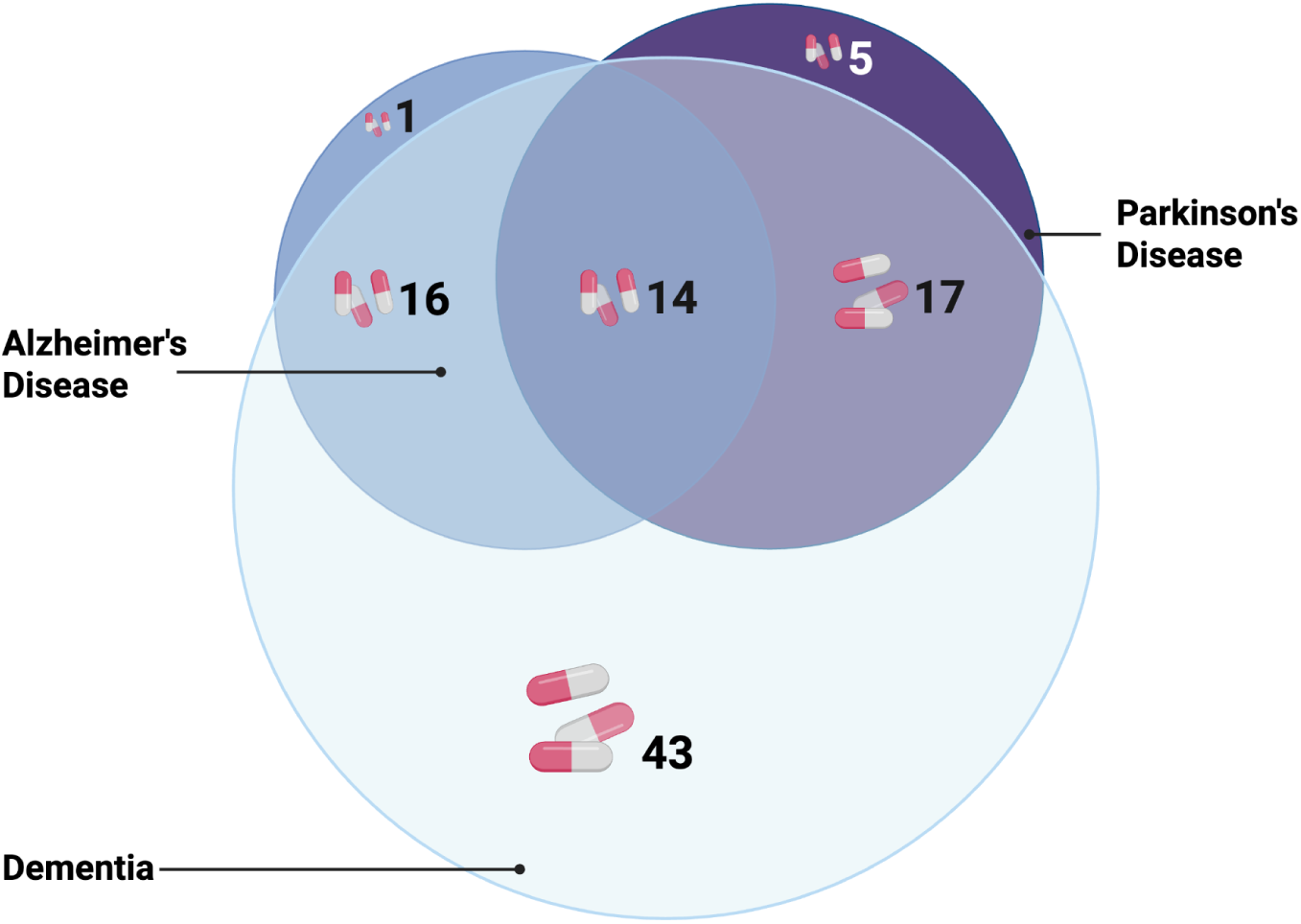
Venn Diagram for 96 medications with replicated associations.

Of the 96 significant medications, we considered 7 to be positive controls: amantadine, donepezil, entacapone, memantine, pramipexole, rivastigmine, and ropinirole, all of which are dementia or PD treatments. Of the remaining 89 medications, 12 were antidepressants, 6 were anticonvulsants, and 5 were antipsychotics. See Table 1 for all significant and replicated medications by medication type.

**Table 1:** Medication type and medication name (generic) associated with risk of NDDs. Positive control* denotes known NDD treatments. The table also lists the medical conditions for which those medications are prescribed, as well as the associated ICD10 codes.

| drug_type | N | medications (generic) | conditions | ICD10 |
| --- | --- | --- | --- | --- |
| antidepressant | 12 | citalopram, duloxetine, escitalopram, mirtazapine, paroxetine, sertraline, trazodone, venlafaxine, amitriptyline, fluoxetine, imipramine, rasagiline | depression, anxiety disorders, OCD, post-traumatic stress disorder, eating disorders, bipolar disorder | F32, F33, F41, F42, F43, F50, F31 |
| positive control | 7 | memantine, rivastigmine, pramipexole, ropinirole, amantadine, donepezil, entacapone | NDD treatments | n/a |
| anticonvulsant | 6 | gabapentin, pregabalin, carbamazepine, lamotrigine, levetiracetam, phenytoin | epilepsy (seizures), nerve pain from shingles, restless leg syndrome, fibromyalgia, | G40, B02, G25, M79 |
| antipsychotic | 5 | quetiapine, haloperidol, olanzapine, prochlorperazine, risperidone | schizophrenia, bipolar disorder, depression | F20, F31, F32, F33 |
| benzodiazepine | 4 | diazepam, lorazepam, temazepam, clonazepam | anxiety disorders, seizures, panic disorder, insomnia | F40, F41, F42, F43, F44, F45, G40, G47, F51 |
| beta blocker | 4 | atenolol, metoprolol, propranolol, sotalol | angina, hypertension, heart rhythm disorders, heart attack, atrial fibrillation, tachycardia | I20, I10, I49, I21, I48, I47 |
| diuretic | 4 | furosemide, bendroflumethiazide, bumetanide, spironolactone | fluid retention (edema) in congestive heart failure, liver disease, kidney disease, hypertension, hypokalemia, nephrotic syndrome | I50, K70, K71, K72, K73, K74, K75, K76, K77, N18, N19, I10, E87, N04 |
| opioid | 4 | codeine, tramadol, morphine, oxycodone | treats symptoms -- pain/coughing | n/a |
| overactive bladder | 4 | solifenacin, tolterodine, mirabegron, trospium | overactive bladder, incontinence, neurogenic detrusor overactivity | N32, N39, N31 |
| proton pump inhibitor | 4 | esomeprazole, pantoprazole, rabeprazole, lansoprazole | gastroesophageal reflux disease; erosive esophagitis | K21, K20, K22 |
| alpha blocker | 3 | alfuzosin, tamsulosin, doxazosin | benign prostatic hyperplasia | N40 |
| anti-diabetic | 3 | glimepiride, rosiglitazone, sitagliptin | type 2 diabetes mellitus | E11 |
| antibiotic | 3 | trimethoprim, erythromycin, nitrofurantoin | bladder infections (UTI), kidney infections, ear infections, dental infections, intestinal infections | N30, N39, N10, H66, K04, K05, A04 |
| calcium channel blocker | 3 | amlodipine, diltiazem, nifedipine | angina, coronary artery disease, hypertension, atrial fibrillation, tachycardia | I20, I25, I10, I11, I12, I13, I15, I16, I48, I47 |
| antacid | 2 | cimetidine, ranitidine | gastroesophageal reflux disease (GERD), stomach ulcer | K21, K25 |
| anti-spasmodic | 2 | oxybutynin, baclofen | overactive bladder, incontinence, night-time urination, spasticity, multiple sclerosis | N32, N39, G81, G35 |
| bisphosphonate | 2 | risedronate, alendronate (alendronic acid) | osteoporosis, Paget's disease | M80, M81, M82, M88 |
| diabetes | 2 | insulin, metformin | diabetes | E10, E11 |
| laxative | 2 | bisacodyl, docusate | constipation | K59 |
| NSAID | 2 | celecoxib, rofecoxib | arthritis, ankylosing spondylitis, menstrual pain, migraine | M06, M13, M15, M19, M45, N94, G43 |
| steroid | 2 | prednisolone, fludrocortisone | Addison's disease, adrenogenital syndrome, arthritis, lupus, psoriasis, ulcerative colitis | E27, E25, M15, M16, M17, M18, M19, M32, L40, K51 |
| ACE inhibitor | 1 | lisinopril | hypertension, congestive heart failure | I10, I11, I12, I13, I15, I16, I50 |
| anti-diarrheal | 1 | loperamide | treats symptoms -- diarrhea | n/a |
| anti-inflammatory | 1 | aspirin | heart attack, angina, stroke | I21, I20, I60, I61, I62, I63, I64, I65, I66, I67, I68, I69 |
| anticoagulant | 1 | warfarin | Embolism and thrombosis | I82 |
| antiemetic & prokinetics | 1 | metoclopramide | gastroesophageal reflux, gastroparesis | K21, K31 |
| antifungal | 1 | nystatin | skin infections caused by yeast | B37 |
| antihistamine | 1 | hydroxyzine | anxiety, hives, contact dermatitis | F40, F41, F42, F43, F44, F45, L50, L25 |
| antiplatelet | 1 | clopidogrel | heart attack, angina | I21, I20 |
| anxiolytic | 1 | buspirone | anxiety | F40, F41, F42, F43, F44, F48 |
| cardiac glycoside | 1 | digoxin | heart failure, atrial fibrillation | I50, I48 |
| cholesterol | 1 | fenofibrate | high cholesterol (hyperlipidemia) | E78 |
| mood stabiliser | 1 | lithium | bipolar disorder | F31 |
| statin | 1 | simvastatin | high cholesterol (hyperlipidemia) | E78 |

The Anticholinergic Cognitive Burden (ACB) score refers to the cumulative impact of taking one or more medications with anticholinergic properties. The ACB score for 28 of these 89 medications was 1 or higher; 11 medications had the highest possible cognitive burden score of 3. An ACB score of 3 or higher has been linked to increased risk of cognitive decline, functional impairment, falls, and mortality in older adults. Higher doses and prolonged use are associated with increased risk of developing dementia.^13–15^ See <u>Table S1</u> for a breakdown of all replicated medications, with ACB burden score available in the “Replicated” tab.

### Adjusting for conditions that increase risk

Although there were many medications that we found to be associated with increased risk of neurodegeneration in the first stage of our analysis, it seemed likely that many of these associations were confounded by underlying conditions, such as depression or hypertension, already known to be risk factors for NDDs. To account for this, we reviewed the significant and replicated medications from our initial analysis using a reputable online source (<u>drugs.com</u>) and recorded the listed conditions for each prescription. We then identified the corresponding ICD10 codes for these conditions and included them as covariates in our follow-up models. The conditions included anxiety disorders, mood disorders, diabetes, epilepsy, hypertension, sleep disorders, and viral infections, among others; see Table 1 for the complete list. As with the medication prescriptions, the ICD10 codes were right censored to only include conditions first diagnosed before NDD diagnosis. We used all ICD10 codes in Table 1 for each regression to account for high rates of comorbidities among older adults. For example, when testing a drug used to treat epilepsy, we also included ICD10 codes for depression and high cholesterol in our analysis.

We re-ran all 480 medication–disease pairings. In UKB, 106 associations passed FDR correction, and 22 of these replicated in AoU. Among the replicated associations, all 7 expected positive controls remained: amantadine, donepezil, entacapone, memantine, pramipexole, rivastigmine, and ropinirole. Rasagiline, a monoamine oxidase inhibitor (MAOI) antidepressant, also replicated in our analysis. However, since it is also used to treat PD, the observed association with dementia likely reflects PD patients with dementia that were included in the dementia cohort. Supporting this, the association with rasagiline was no longer significant after excluding PD patients from the DEM group. See <u>Table S2</u> for all results.

Removing the positive controls leaves 10 significant and replicated associations – 2 for AD, 6 for DEM, and 2 for PD – with a total of 9 unique medications. Of those 9 unique medications, 3 antidepressants, 2 anticonvulsants, and 2 antipsychotics remained significant and replicated, as well as 1 benzodiazepine and 1 beta blocker. See Table 2 for all significant and replicated pairings and <u>Table S2</u> for all results.

**Table 2:** Medication and NDD associations persist after adjusting for known ICD10 codes in our two-stage design. The pairs with a * were no longer replicated when *APOE*-ε4 was added to the analysis.

| NDD | MED | DESCRIPTION | acb | COHORT | HR | ci_min | ci_max | N_pairs | N | P_CORR |
| --- | --- | --- | --- | --- | --- | --- | --- | --- | --- | --- |
| AD | quetiapine | antipsychotic | 3 | UKB | 5.70 | 2.86 | 11.36 | 11 | 420 | 7.62E-06 |
|  |  |  |  | AoU | 2.76 | 1.77 | 4.31 | 26 | 1193 | 1.32E-04 |
|  | sertraline* | antidepressant | 0 | UKB | 3.05 | 2.30 | 4.04 | 72 | 7221 | 9.58E-14 |
|  |  |  |  | AoU* | 1.59 | 1.18 | 2.14 | 64 | 4807 | 2.32E-02 |
| DEM | duloxetine | antidepressant | 0 | UKB | 1.76 | 1.28 | 2.41 | 44 | 1696 | 2.96E-03 |
|  |  |  |  | AoU | 1.54 | 1.31 | 1.83 | 189 | 3586 | 6.65E-06 |
|  | gabapentin | anticonvulsant | 0 | UKB | 1.38 | 1.14 | 1.67 | 136 | 8691 | 5.27E-03 |
|  |  |  |  | AoU | 1.30 | 1.15 | 1.47 | 545 | 13655 | 2.42E-04 |
|  | mirtazapine | antidepressant | 0 | UKB | 3.14 | 2.60 | 3.80 | 155 | 4420 | 6.23E-31 |
|  |  |  |  | AoU | 1.49 | 1.23 | 1.80 | 150 | 1873 | 6.83E-04 |
|  | pregabalin | anticonvulsant | 0 | UKB | 1.73 | 1.37 | 2.19 | 84 | 4055 | 3.98E-05 |
|  |  |  |  | AoU | 1.76 | 1.47 | 2.12 | 142 | 2388 | 3.99E-08 |
|  | quetiapine | antipsychotic | 3 | UKB | 6.31 | 4.34 | 9.18 | 43 | 452 | 8.70E-21 |
|  |  |  |  | AoU | 1.73 | 1.41 | 2.13 | 144 | 1311 | 4.34E-06 |
|  | risperidone | antipsychotic | 1 | UKB | 7.24 | 5.05 | 10.38 | 41 | 331 | 7.91E-26 |
|  |  |  |  | AoU | 2.06 | 1.49 | 2.84 | 50 | 469 | 1.94E-04 |
| PD | clonazepam | benzodiazepine | 0 | UKB | 12.02 | 6.50 | 22.22 | 12 | 326 | 3.03E-14 |
|  |  |  |  | AoU | 2.14 | 1.63 | 2.81 | 74 | 2450 | 9.38E-07 |
|  | propranolol | beta blocker | 0 | UKB | 2.65 | 1.94 | 3.63 | 50 | 9226 | 1.37E-08 |
|  |  |  |  | AoU | 3.29 | 2.41 | 4.48 | 51 | 1578 | 1.48E-12 |

We also added *APOE*-ε4 status as a covariate to further correct for and serve as a proxy for unrecorded dementia or MCI that can be present in population-style biobanks. When we added *APOE*-ε4 status to our analysis for these 10 pairings, 9 remained significant and replicated. Sertraline was no longer replicated for AD in AoU. See <u>Table S2</u> for all results.

Including potentially comorbid ICD10 codes as covariates provided additional insight to our models. In UKB, we ran 480 Cox regressions. 4 ICD10 codes were significant in more models than traditional demographic covariates such as age, sex, or the Townsend deprivation index: I67 (Other cerebrovascular diseases, including strokes and aneurysms), G40 (Epilepsy), F32 (Depressive episode), and N39 (Other disorders of the urinary system, including urinary tract infections). These ICD10 codes remained significant in more models than age, sex, or Townsend index even after adjusting for APOE genotype (e3/e4 or e4/e4). In AoU, we had enough data to run 433 regressions. In addition to the ICD10 codes identified in UKB, several others were significant in more models than age or sex: G47 (Sleep disorders), I63 (Cerebral infarction, or stroke), M79 (Other soft tissue disorders, including fibromyalgia), and F31 (Bipolar disorder). (Townsend deprivation index was not available in AoU) See <u>Table S2</u> “significant_covaries” tab for a list of all covariates that were significant in at least 100 models.

For the significant associations in Table 2, we also ran an interaction analysis for each drug and *APOE*-ε4 carrier status. There were no drugs for which the interaction term was significant and replicated across datasets after multiple test corrections.

### Associations between number of medication exposures and neurodegeneration

We next aimed to account for the number of times an individual was exposed to a medication. While dosage information was too sparse and inconsistently recorded in the EHR to be useful, we decided to use the number of times an individual had a prescription date in their EHR as a proxy for exposure. We used quartiles to divide the number of prescriptions per individual into low, medium, and high users for each medication; i.e. low users were in the 1st quartile; medium users in the 2nd and 3rd, and high users were in the 4th quartile. We also used the ICD10 codes described above as covariates.

8 medications (excluding positive controls) were significant and replicated after dividing the data into low, medium, and high users. Adjusting for *APOE*-ε4 carrier status added one more significant medication (atorvastatin) to our table. (Table 3) All medications remained significant at the reported usage levels, regardless of *APOE*-ε4 status, except for those marked with an asterisk (*), which lost significance when *APOE* was included as a covariate. In contrast, cohorts labeled with the suffix “-*APOE*” became significant only after *APOE* status was added to the analysis.

**Table 3:** This table shows NDD/medication pairings that remained significant and replicated at different levels of exposure in both UKB and AoU. Groupings with a * were no longer replicated when *APOE*-ε4 was added to the analysis.Groupings with “-*APOE*” were only significant when *APOE* was included in the analysis. A HR < 1 suggests a protective association.

| NDD | PRIOR | description | acb | USE | COHORT | HR | ci_min | ci_max | N_pairs | N | P_CORR |
| --- | --- | --- | --- | --- | --- | --- | --- | --- | --- | --- | --- |
| AD | aspirin | anti-inflammatory | 0 | high | UKB | 2.51 | 1.81 | 3.48 | 69 | 6973 | 5.75E-07 |
|  |  |  |  |  | AoU | 1.57 | 1.16 | 2.11 | 101 | 6457 | 4.40E-02 |
| DEM | atorvastatin | statin | 0 | low | UKB | 0.67 | 0.50 | 0.89 | 52 | 7003 | 3.25E-02 |
|  |  |  |  | med | AoU | 0.78 | 0.67 | 0.92 | 217 | 9606 | 4.16E-02 |
|  |  |  |  |  | UKB - *APOE* | 0.74 | 0.61 | 0.89 | 135 | 13720 | 1.29E-02 |
|  | gabapentin | anticonvulsant | 0 | med | UKB | 1.51 | 1.15 | 1.99 | 58 | 3644 | 1.98E-02 |
|  |  |  |  | high | UKB | 1.68 | 1.26 | 2.23 | 56 | 2265 | 3.71E-03 |
|  |  |  |  |  | AoU | 1.60 | 1.35 | 1.89 | 235 | 3467 | 1.55E-06 |
|  | mirtazapine | antidepressant | 0 | low | UKB | 3.07 | 2.15 | 4.37 | 33 | 1225 | 1.26E-08 |
|  |  |  |  |  | AoU | 1.84 | 1.29 | 2.61 | 35 | 519 | 1.38E-02 |
|  |  |  |  | med | UKB | 2.79 | 2.13 | 3.67 | 61 | 2084 | 3.59E-12 |
|  |  |  |  | high | UKB | 3.74 | 2.81 | 4.98 | 61 | 1111 | 8.25E-18 |
|  |  |  |  |  | AoU - APOE | 2.03 | 1.48 | 2.78 | 46 | 407 | 3.76E-04 |
|  | pregabalin | anticonvulsant | 0 | med | UKB | 1.89 | 1.38 | 2.60 | 42 | 1955 | 9.08E-04 |
|  |  |  |  |  | AoU* | 1.67 | 1.25 | 2.25 | 49 | 909 | 1.30E-02 |
|  |  |  |  | high | UKB | 1.80 | 1.21 | 2.67 | 28 | 1022 | 2.49E-02 |
|  |  |  |  |  | AoU | 2.23 | 1.68 | 2.96 | 59 | 679 | 1.37E-06 |
|  | quetiapine | antipsychotic | 3 | low | UKB | 10.30 | 5.88 | 18.04 | 14 | 125 | 1.11E-14 |
|  |  |  |  | med | UKB | 5.33 | 3.24 | 8.77 | 21 | 209 | 1.05E-09 |
|  |  |  |  |  | AoU | 1.80 | 1.37 | 2.36 | 73 | 626 | 6.89E-04 |
|  |  |  |  | high | UKB | 4.75 | 2.21 | 10.19 | 8 | 118 | 6.94E-04 |
|  |  |  |  |  | AoU - APOE | 2.28 | 1.58 | 3.30 | 37 | 270 | 3.12E-04 |
|  | risperidone | antipsychotic | 1 | low | UKB | 6.37 | 3.44 | 11.80 | 13 | 100 | 7.35E-08 |
|  |  |  |  |  | AoU | 2.68 | 1.75 | 4.11 | 23 | 173 | 1.99E-04 |
|  |  |  |  | med | UKB | 8.79 | 5.53 | 13.98 | 21 | 146 | 1.72E-18 |
|  |  |  |  | high | UKB | 5.23 | 2.35 | 11.65 | 7 | 85 | 5.91E-04 |
| PD | propranolol | beta blocker | 0 | low | UKB | 2.10 | 1.22 | 3.62 | 14 | 3330 | 4.15E-02 |
|  |  |  |  |  | AoU | 3.07 | 1.63 | 5.79 | 10 | 449 | 1.18E-02 |
|  |  |  |  | med | UKB | 3.04 | 1.93 | 4.79 | 21 | 3482 | 2.30E-05 |
|  |  |  |  |  | AoU | 2.77 | 1.76 | 4.37 | 23 | 710 | 3.50E-04 |
|  |  |  |  | high | UKB | 2.85 | 1.67 | 4.85 | 15 | 2414 | 1.21E-03 |
|  |  |  |  |  | AoU | 4.40 | 2.71 | 7.15 | 18 | 419 | 1.07E-07 |

7 of these medications were significant for high users in at least one cohort; 7 of these medications were also significant for medium usage. 5 medications remained significant at low usage. One medication, atorvastatin, had a protective effect at low or medium usage, but this effect was no longer significant at high usage. Two medications, aspirin and atorvastatin, did not appear in Table 2, which looked at any level of exposure. Of these 8 medications, there were 2 anticonvulsants and 2 antipsychotics. Only 1 antidepressant, mirtazapine, remained significant. See Table 3 for complete replicated results with usage level and <u>Table S3</u> for all results.

### First medication exposure ten or more years before tenure

To assess how medication taken years prior to NDD diagnosis impacted risk, we repeated our analysis, restricting it to medications first taken 10 or more years before the endpoint (defined as NDD diagnosis, death, or end of study follow-up). Several positive controls replicated again at this time point, including the dopamine agonists pramipexole and ropinirole. However, these medications can be used to treat PD and Restless Legs Syndrome (RLS); they did not remain significant when we remove PD cases with dementia from our dementia cases. See <u>Table S4</u> for complete results.

After removing positive controls, 4 medication/NDD pairings were significant and replicated, 3 for DEM and 1 for PD. All medications had HR < 1, which suggests a protective association (see medications in bold in Table 4). 2 of the significant medications were antibiotics: amoxicillin, which showed a protective effect for both DEM and PD, and ciprofloxacin, which was associated with a protective effect for DEM. Neither medication was significant in our analyses when considering exposure at any time point (Table 2). The final medication that remained significant 10 years before NDD diagnoses was omeprazole, a proton pump inhibitor, which was associated with a protective effect for DEM.

**Table 4:** This table shows NDD/medication pairings replicated by medication name in bold; the others replicated by medication type. These pairings remain significant and replicated in both cohorts with a first exposure occurring ten or more years before tenure. The cohorts with a * were no longer replicated when *APOE*-ε4 was added to the analysis.

| medication_type | NDD | medication | acb | COHORT | LAG | HR | ci_min | ci_max | P_VAL | N_pairs | N | P_CORR |
| --- | --- | --- | --- | --- | --- | --- | --- | --- | --- | --- | --- | --- |
| antibiotic | DEM | **amoxicillin** | 0 | UKB | 10+ | 0.62 | 0.54 | 0.71 | 1.88E-12 | 324 | 49752 | 1.16E-10 |
|  |  |  |  | AoU | 10+ | 0.68 | 0.58 | 0.81 | 9.55E-06 | 162 | 7611 | 4.97E-04 |
|  |  | azithromycin | 0 | AoU | 10+ | 0.70 | 0.59 | 0.83 | 4.06E-05 | 165 | 6825 | 1.19E-03 |
|  |  | **ciprofloxacin** | 0 | UKB* | 10+ | 0.67 | 0.51 | 0.87 | 3.13E-03 | 60 | 8036 | 3.53E-02 |
|  |  |  |  | AoU | 10+ | 0.60 | 0.50 | 0.72 | 1.33E-07 | 131 | 5811 | 1.83E-05 |
|  |  | co-amoxiclav | 0 | UKB | 10+ | 0.68 | 0.54 | 0.86 | 1.38E-03 | 76 | 11804 | 2.02E-02 |
|  |  | flucloxacillin | 0 | UKB | 10+ | 0.77 | 0.65 | 0.90 | 1.24E-03 | 179 | 24926 | 1.92E-02 |
|  |  | metronidazole | 0 | AoU* | 10+ | 0.60 | 0.45 | 0.80 | 6.11E-04 | 56 | 1926 | 1.29E-02 |
|  |  | nitrofurantoin | 0 | AoU | 10+ | 0.41 | 0.27 | 0.62 | 2.03E-05 | 28 | 1327 | 7.96E-04 |
|  |  | penicillin | 0 | UKB | 10+ | 0.48 | 0.31 | 0.75 | 1.04E-03 | 21 | 5257 | 1.73E-02 |
|  |  | trimethoprim | 3 | UKB | 10+ | 0.67 | 0.55 | 0.81 | 5.14E-05 | 127 | 19185 | 1.42E-03 |
|  | PD | **amoxicillin** | 0 | UKB | 10+ | 0.57 | 0.44 | 0.73 | 8.08E-06 | 87 | 49541 | 2.86E-04 |
|  |  |  |  | AoU* | 10+ | 0.65 | 0.49 | 0.85 | 1.88E-03 | 62 | 7511 | 2.53E-02 |
| high blood pressure treatment | DEM | ramipril (ACE inhibitor) | 0 | UKB | 10+ | 0.64 | 0.49 | 0.82 | 4.91E-04 | 69 | 7559 | 9.36E-03 |
|  |  | lisinopril (ACE inhibitor) | 0 | AoU | 10+ | 0.74 | 0.62 | 0.87 | 4.23E-04 | 175 | 5532 | 9.67E-03 |
|  |  | perindopril (ACE antihypertensive) | 0 | UKB | 10+ | 0.49 | 0.31 | 0.79 | 3.07E-03 | 18 | 2388 | 3.53E-02 |
|  |  | losartan (ARB antihypertensive) | 0 | AoU | 10+ | 0.48 | 0.35 | 0.67 | 1.01E-05 | 45 | 1642 | 4.97E-04 |
|  |  | bisoprolol (beta blocker) | 0 | UKB | 10+ | 0.55 | 0.37 | 0.83 | 4.56E-03 | 25 | 2448 | 4.92E-02 |
|  |  | metoprolol (beta blocker) | 1 | AoU | 10+ | 0.64 | 0.52 | 0.78 | 8.54E-06 | 127 | 3462 | 4.97E-04 |
| proton pump inhibitor | DEM | omeprazole | 0 | UKB | 10+ | 0.60 | 0.48 | 0.74 | 1.29E-06 | 104 | 15020 | 5.35E-05 |
|  |  |  |  | AoU | 10+ | 0.70 | 0.59 | 0.83 | 4.34E-05 | 179 | 5981 | 1.19E-03 |

The significant inverse effect on diseases seen 3 times for antibiotics led us to consider whether other antibiotics might have shown significance but failed to replicate due to differences in prescribing patterns between countries. For instance, bisoprolol is one of the most commonly prescribed beta blockers in the UK, while metoprolol is more frequently prescribed in the USA.^16,17^ Thus, we expanded Table 4 to group medications by type, highlighting those that replicated across cohorts with a first exposure occurring 10 or more years before the endpoint.

After expanding by drug type, seven other antibiotics were significant in at least one cohort (Table 4). These were evenly divided between AoU and UKB, and all were protective, with HR ranging from 0.41 for nitrofurantoin in AoU to 0.77 for flucloxacillin in UKB. Grouping by medication type, six medications that treat high blood pressure were also significant and protective with HRs ranging from 0.48 for losartan in AoU to 0.74 for lisinopril in AoU. Interestingly, different types of high blood pressure treatments (ACE inhibitors, beta blockers, and one ARB) showed similar types of protective effects. We did not find any additional significant medications in the proton pump inhibitor group besides omeprazole.

## Discussion

Our analysis reinforces existing evidence that certain high-risk medications are associated with NDD phenotypes, while also identifying novel potential medication–disease interactions. We demonstrate the value of incorporating clinical diagnosis data alongside prescription records in risk models, as medication associations often serve as proxies for underlying conditions already known to increase NDD risk.

### Medications that may increase risk of developing an NDD

Although our initial analysis identified 96 medications associated with an increased risk of developing an NDD, adjusting for related conditions using ICD10 codes substantially reduced the number of significant associations. After this adjustment, only 10 medications remained linked to increased risk, including 3 antidepressants, 2 anticonvulsants, 2 antipsychotics, 1 benzodiazepine, 1 beta blocker, and 1 anti-inflammatory drug.

### Antidepressants

Initially, 12 antidepressants were associated with an increased risk of developing a NDD; after adjusting for related conditions including depression, anxiety disorders, OCD, PTSD, eating disorders, and bipolar disorder, only 3 medications remained significant: sertraline, duloxetine, and mirtazapine. Sertraline no longer replicated when we adjusted for *APOE*-ε4 status.

Sertraline is a selective serotonin reuptake inhibitor (SSRI), which is the most commonly prescribed class of antidepressants. When *APOE*-ε4 was added to our analysis, the sertraline association no longer replicated. This may reflect early prescribing in individuals with MCI who were not yet diagnosed with AD or DEM, as depression is a common comorbidity for MCI. We did not adjust directly for MCI in our analyses, but *APOE*-ε4 status likely acted as a proxy for individuals with both depression and early cognitive decline, as *APOE* is a known genetic risk factor for AD and dementia and has also been linked to depression.^18^ Notably, some research suggests that long-term SSRI use may slow the progression from MCI to AD.^19^ While this is only an association, it is worth noting that other SSRIs, such as fluoxetine and paroxetine, did not show a significant association with increased risk after adjusting for related conditions.

The results of past studies looking at duloxetine (a serotonin-noradrenaline reuptake inhibitor (SNRI)) and NDDs have shown inconclusive results.^19,20^ In our study, it was still associated with dementia, even after adjusting for risk conditions and *APOE*-ε4 status.

Mirtazapine is a tetracyclic antidepressant, sometimes referred to as an atypical antidepressant. In the past, mirtazapine was often prescribed for agitation in people with dementia, however recent studies suggest that mirtazapine is not effective for agitation and is associated with increased mortality, so it is no longer recommended for this population.^21^

### Anticonvulsants

Previous studies have reported that the use of gabapentin and pregabalin is linked to an increased risk of developing dementia in epilepsy management and among older adults with previously normal cognition, however, epilepsy itself has also been associated with dementia.^22,23^ A 2023 study looking at gabapentin or pregabalin and DEM found a HR of 1.45 for these medications, similar to the HR we found for these medications in our analysis (HR 1.31 in UKB for gabapentin to HR 1.77 for pregabalin in AoU).^24^ Despite potential risks for cognitive decline, gabapentin has also been explored as a treatment for behavioral symptoms of dementia in older adults, which may affect results in the case of currently undiagnosed dementia cases present in our datasets.^25^ In the United States, gabapentin is also commonly prescribed off-label to treat nerve pain in individuals with multiple sclerosis (MS);^26^ however, we did not adjust for MS in our analysis. While evidence remains conflicting for these drugs given the complexity of the involved conditions, it may be wise to prescribe these with care, especially at high dosages in older adults at risk for cognitive decline, as suggested by previous studies.^27,28^ While this is only an association, there were other anticonvulsants in our study where the association with NDDs did not remain significant (carbamazepine, lamotrigine, levetiracetam, phenytoin).

### Antipsychotics

There were 5 antipsychotic medications that were associated with an increased risk of developing an NDD in our initial analysis; after adjusting for a range of psychiatric comorbidities including schizophrenia, only 2 medications remained significant: quetiapine and risperidone.

These medications are commonly prescribed off-label to manage agitation and aggression in patients with AD or other dementias, however quetiapine has been associated with greater cognitive decline compared to placebo when treating behavioral symptoms in dementia, and risperidone is not approved in the US for this use.^29^ Risperidone is approved for treatment of schizophrenia, which is a condition associated with increased risk of dementia,^30^ but it is worth noting that adjusting for schizophrenia and other psychiatric conditions resulted in 3 (haloperidol, olanzapine, and prochlorperazine) of the initially significant antipsychotics to lose their association with dementia, while the associations for quetiapine and risperidone remained.

### Benzodiazepines

4 benzodiazepines were initially associated with later development of an NDD; this reduced to 1 medication, clonazepam, when we adjusted for conditions including anxiety disorders and seizure disorders. Often used in treating REM sleep-related behavior disorder (RBD),^31^ this association is once again likely a proxy for early phases of PD as RBD was unavailable in our datasets to use as an adjustment in the models.

### Beta blockers

Our initial analysis nominated 4 beta blockers that increased the risk of developing an NDD; this fell to 1 medication when we adjusted for related ICD10 codes. The remaining association was for propranolol and PD, a relationship that has been investigated before in previous literature.^32,33^ While results are conflicting, propranolol is often used to treat tremors unrelated to or pre-onset of PD, so it is possible that the association we see in our study is simply a proxy for early phases of PD.

### Anti-inflammatory

The last medication that our analysis flagged as increasing risk of developing an NDD after adjusting for comorbidities was aspirin, but only at high levels (Table 3). While we did adjust for the ICD10 code for heart disease, we did not account for the severity of the condition. As aspirin may be prescribed for a wide variety of health issues, this association is potentially a proxy for additional health issues or coronary heart disease, which is associated with dementia.^34^

### Medications that may decrease risk of developing an NDD

One of the most interesting findings from our study was that 3 classes of medications showed significant protective associations after adjusting for comorbid conditions – but only when first taken at least ten or more years prior to the study endpoint (defined as death, end of follow-up, or NDD diagnosis). These drug classes included antibiotics, high blood pressure treatments, and proton pump inhibitors.

### Antibiotics

The two antibiotics that showed a replicated protective effect were amoxicillin and ciprofloxacin. Amoxicillin is commonly used to treat dental or sinus infections, and ciprofloxacin is often prescribed for urinary tract infections (UTI). Ciprofloxacin is a fluoroquinolone antibiotic and is known to cross the blood-brain barrier.^35^ Previous studies have also found associations between chronic periodontitis and dementia.^36^ ^37^ UTIs are known to cause confusion, delirium, and worsening of cognitive impairment in older adults. Our models were adjusted for pulpitis (inflammation inside of a tooth), periapical abscess (infection at the tooth root), and UTIs.

Very high doses of amoxicillin have been shown to be neurotoxic in previous literature, while others suggest there are neuroprotective effects for AD related to moderate dosing of certain β-lactam antibiotics.^38,39^ Ciprofloxacin has been targeted as a potential therapeutic for ALS and AD, while also being flagged as potentially increasing risk for peripheral neuropathy and presenting some central nervous system side effects.^40–43^ These conflicting results, including some results from animal models, suggest a need for further mechanistic follow-up. These associations may be proxies for bacterial or other infections connected to disease etiology.^44^ Prescription records from over 10 years ago may also reflect individuals in generally better health, particularly those who engage more regularly with healthcare services and seek treatment for minor infections throughout their lives. Further research is needed to better understand the potential role of antibiotics in reducing the risk of AD and dementia.

### Medications that lower blood pressure

Previous studies have found that controlling blood pressure reduced the risk of mild cognitive impairment.^12^ A more recent study from the Netherlands found that angiotensin II receptor blockers (ARBs), beta blockers, and diuretics all lowered the risk of dementia.^45^ This study adjusted their models for heart related events, including myocardial infarction, stroke, and congestive heart failure. Our study yielded similar findings, showing that the ARB losartan, along with two beta blockers (bisoprolol and metoprolol), were associated with a reduced risk of NDDs. We also identified several ACE inhibitors associated with lower risk. However, protective associations for all antihypertensives were replicated at the drug class level rather than for specific individual medications.

Statins are primarily prescribed to lower cholesterol levels, but they may also help lower blood pressure. In our study, atorvastatin was associated with a reduced risk of developing dementia; however, this association replicated only at medium usage levels (Table 3). One other note of interest is that the association in UKB was only significant after including *APOE*-ε4 status, supporting some previous literature that suggests statins may be more effective for carriers of *APOE*-ε4.^9^

### Proton pump inhibitors

Research on the association between proton pump inhibitor (PPI) use and dementia risk has yielded mixed results. One study by Gomm et al. reported an increased risk of dementia with PPI use, but their analysis adjusted for only 14 ICD10 codes, whereas our model included over 70.⁴⁴ Another study with similar findings adjusted for conditions such as stroke, heart disease, diabetes, hypertension, and hyperlipidemia, similar to those considered by Gomm et al.⁴⁵ However, neither study accounted for other important comorbidities like epilepsy, depression, urinary tract infections, or sleep disorders, which, in our systematic and hypothesis-free analysis, emerged as more consistently significant covariates than age or sex.

Depression and gastrointestinal issues are known to be linked, raising the possibility that studies examining PPIs may be capturing underlying depression risk rather than a direct effect of the medication⁴⁶. In our own analysis, PPIs were initially associated with increased dementia risk before adjusting for ICD10 codes (<u>Table S1</u>). However, other studies that also included depression as a covariate, including a large study from Wales, and found a protective association for PPIs.⁴⁷ The HR reported in that study (0.67) closely aligns with the results from our ICD10–adjusted models (UKB: 0.60, AoU: 0.70; <u>Table S4</u>).

For all of these medications, further research is needed to disentangle the complex interactions with NDD risk. Our findings highlight the importance of adjusting for a broad range of risk factors in association studies, not just the conditions for which a medication is prescribed. Overall, our results are reassuring: the vast majority of medications were not associated with an increased risk of developing an NDD. In addition, our study suggests that effectively treating common conditions such as infections, high blood pressure, and gastrointestinal issues may be linked to a reduced risk of developing NDDs later in life.

### Limitations

This study has several limitations. One is that the medication data in UKB only extends to 2018, meaning that we lack medication records for many NDD cases diagnosed after this period. Due to this, we had to exclude all cases diagnosed after 2018 in UKB, reducing our sample size and ability to test more recently diagnosed cases in this dataset. We were also unable to account for dose and duration of time an individual took a medication – or if they actually picked up and took their prescription. Additionally, because we rely on biobank data for this study, there is always the possibility of inaccuracies in diagnoses and gaps in recorded medical history. The medication data in UKB is sparse before 1999, limiting our ability to assess long-term medication use for some individuals accurately. Another potential limitation is that, while we attempted to add relevant covariates to explain the associations seen between certain medications and NDD risk, we may not have accounted for all relevant covariates, potentially leading to residual confounding effects. We were also unable to replicate all of our discovery results in AoU due to smaller NDD sample sizes in that dataset, as some medications were recorded for fewer than 10 NDD cases. Finally, differences in drug approvals and healthcare practices between the US and UK may have influenced our findings. Additionally, it is important to note that the current analysis does not address the effect of medication exposure on NDDs compared to the risk associated with cessation of treatment for other disorders, a balance in risk that should be thoughtfully addressed in a clinical context.

A potentially interesting future direction for this study could be to investigate drug metabolism, for example, assessing the effect of interactions in CYP drug metabolizer genes and medication on NDD risk. It would also be interesting to investigate drug interactions in patients who were prescribed more than one medication.

### Conclusions

Our study identified and replicated both risk and protective associations with NDD risk across two large biobanks, using a hypothesis-free, open science approach. By leveraging open-access data and code, we provide a foundation to enable researchers to conduct further investigation in additional datasets to explore underlying mechanisms and interactions in greater depth. Our findings underscore the importance of adjusting for comorbidities that may independently influence risk. While these results are observational and do not establish causality, they highlight associations that may have meaningful, translational relevance for future research and clinical practice.

## Supplementary Tables

**<u>Table S1</u>**: Cox baseline any exposure for UKB and AoU – all results

**<u>Table S2</u>**: Cox with ICD10 codes and APOE for UKB and AoU – all results

**<u>Table S3</u>**: Cox with different levels of exposure (low, medium, and high) – all results

**<u>Table S4</u>**: Cox with first exposure ten years before tenure – all results

**<u>Table S5</u>**: Medication short name table

## Methods

This study uses data from two biobanks: the UK Biobank (UKB) and All of Us (AoU) in a two-stage design. UKB was chosen as the larger discovery set and AoU as the replication dataset.

### Medication Data in UKB

UKB medication data came from Data-Field 42039: GP2 prescription records. This field contains 56 million rows of data from four different sources: England (TPP), Wales, England (Vision) and Scotland. Read2 and bnf codes were provided for some (but not all) of the medications. Staff at UKB confirmed there was no common data column to link medications from all four sources.

Since there was no common medication ID, we created our own method to use the human readable medication description to extract common medication types. Using the list of commonly found medications in the National Heath System as a starting point, we manually created a spreadsheet that allowed us to group medications with the same active ingredient together. <u>Table S5</u> For each medication, we created a “cleaned_med” name that we used to assign a “short_name”, which was the generic version of that medication.

For example, Atenix, Lustral, Zoloft, as well as the generic sertraline, would all be given the “short_name” of “sertraline”. See example below:

| medication | cleaned_med | type | short_name | description | notes |
| --- | --- | --- | --- | --- | --- |
| Atenix | atenix | brand | sertraline | antidepressant | SSRI |
| Lustral | lustral | brand | sertraline | antidepressant | SSRI |
| Sertraline | sertraline | generic | sertraline | antidepressant | SSRI |
| Zoloft | zoloft | brand | sertraline | antidepressant | SSRI |

For sertraline, this method allowed us to group 55 different “medication names” with the same active ingredient together. A few examples of the different types of “medication names” that would be grouped together: Sertraline 50mg tablets, SERTRALINE 50mg tablets, LUSTRAL 50mg tablets, Atenix 25 tablets (Ashbourne Pharmaceuticals Ltd), etc. This allowed us to greatly increase the number of prescriptions we had for each medication type. We then looked at associations between the “short_name” (aka the generic name) for each medication and each of our NDDs. For this study we did not differentiate between different dosages of the same medication.

We also decided to omit vitamins, supplements, nasal sprays, inhalers, eye/ear drops, lotions, hormones, and vaccines from our study due to regional variation in over the counter medication naming. We also tried to omit combination medications. With this method, we identified 1292 unique medication names in UKB and 100 keywords used to omit medications. 336 medications were omitted because they fell into the above excluded categories. This method accounted for 99% of the medication data available in UKB.

We also only included data from 1999 or later, as data before 1999 was very sparse. This left us with 537 medications (short_names) with data, spanning the time period from 1999-2017. After restricting to medications with at least 10 overlapping NDD–medication pairings, we were able to test a total of 224 medications. An advantage of this approach is that we could group medications with the same active ingredients together. It is also transferable to any medication database that contains a human readable name, making the grouping process easier to understand. The tables we used for these groupings can be found in <u>Table S5</u> and code for the data cleaning can be found on our GitHub.

Medication data is only available for 45% of the entire UKB cohort, leaving us with 222,073 participants with available medication data. NDD data and control cohorts were pulled from this subsample of the UKB, to ensure we only had participants who had recorded medication data available.

### Medication Data in All of Us

AoU was used as a replication dataset. Of the 224 medications tested, we were able to pull data for 187 in AoU. We added medication data using the All of Us Workbench and manually added all 187 medication names to create a concept set. While AoU’s medication data was cleaner than UKBs and included RxNorm encodings for each prescription, to be consistent we decided to use the same method we used in UKB for grouping and processing different medication types. This meant that medications with descriptions such as “sertraline”, “sertraline 50 mg oral tablet”, and “sertraline 100 mg oral tablet [zoloft]” would all be grouped together as we did with UKB. A few medications had different names in the UK and in the US – in those cases both names were included in our results.

At the time of our analysis, medication data was available for 334,162 participants in AoU. NDD data and control cohorts were pulled from this subsample of AoU, to ensure we only had participants who had recorded medication data available. As in UKB, we only included medication data from 1999 or later.

### UKB Cohort Formation

The neurological phenotype for AD was obtained from Data-Field 42040: Date of Alzheimer’s disease report. This is an algorithmically-defined outcome in UKB. Individuals who also had an ALS, FTD, PD, PSP, or MSA diagnosis were removed. For PD, the phenotype was obtained from Data-Field 42032: Date of Parkinson’s disease report. This is an algorithmically-defined outcome in UKB. Individuals with any dementia code, as well as ALS, PSP, or MSA were removed. For DEM, the phenotype was obtained from Data-Field 42018: Date of all cause dementia report. This is an algorithmically-defined outcome in UKB. Comorbidities were not removed for the all-cause dementia (DEM) phenotype; a DEM phenotype includes patients with AD and includes patients with PD who also have dementia. Only incident cases (cases that occurred during the course of the study period) were included.

The control cohort was created by excluding 24 ICD10 codes for NDDs or related conditions. See our GitHub for the entire list. Controls were selected with a minimum current age of 60 for better age matching to our case groups and to exclude any younger controls who may develop an NDD in the future.

Our analysis was limited to individuals of European ancestry. Additionally, because medication data was only available through the end of 2017, we restricted NDD diagnoses to the same time frame. As a result, the number of NDD cases included in our study was substantially smaller than the total number of NDD cases in the full UKB cohort.

### AoU Cohort Formation

The neurological phenotype for AD was obtained using the AoU concept ID 378419. Individuals who also had an ALS, FTD, PD, PSP, or MSA diagnosis were removed. Individuals with “Mixed dementia”, “Early onset”, and “presenile onset” were also excluded. For PD, the phenotype was obtained from AoU concept ID 381270. Individuals with any dementia code, as well as ALS, PSP, or MSA, or vascular dementia were excluded. Individuals with “Young onset” were also excluded. For DEM, the phenotype was obtained from AoU concept ID 4182210. Individuals with “Postconcussion syndrome” and “presenile onset” were also excluded, as they did not match cohort formation in UKB. Comorbidities were not removed for the all-cause dementia (DEM) phenotype. Only incident cases (cases that occurred during the course of the study period) were included.

The control cohort was constructed by excluding the same diagnostic codes and applying the same criteria used in the UK Biobank. Due to sample size constraints, we again limited our analysis to individuals of European ancestry. Both medication and NDD diagnosis data extended through 2023, and we included all available data up to that point.

### Statistical methods

Using time series data for medication exposure and diagnosis dates, we used Cox proportional hazards models implemented in the Python package lifelines 0.26.4 to test the association between medication exposure and NDD diagnosis during the study period, adjusting for age and sex (and Townsend deprivation index in UKB only). The results for each disease underwent Benjamini Hochberg false discovery rate adjustment to adjust P values for multiple testing.

The analysis was later rerun, using 88 ICD10 codes to account for potential confounding by other conditions. See Table 1 for a complete list of ICD10 codes. Codes were only included in our analysis if at least 20 people in our dataset had that diagnosis.

Next, for each medication, we divided each participant into low, medium, or high users, using the number of prescriptions that were recorded in their EHR. These groupings were created by quartile for each medication (low = 1st quartile, medium = 2nd and 3rd quartiles, high = 4th quartile). We then reran the analysis using these three groupings in our model. Finally, we ran the analysis using a minimum ten year lag between starting medication use and tenure (death, diagnosis of an NDD, or end of study).

*APOE*-ε4 dosage was one-hot-encoded for ‘e3/e4’ or ‘e4/e4’. Genetic data was not available for all participants; when not available, participants were removed from the analysis, resulting in a slightly smaller sample size when *APOE*-ε4 dosage was included.

### Data Availability

Data for this study is all open source. The discovery phase of analysis was carried out in the UK BioBank’s Research Analysis Platform (RAP) and can be accessed via https://www.ukbiobank.ac.uk/enable-your-research/research-analysis-platform. The replication phase of analysis was carried out using the AllOfUs Study’s WorkBench Platform and can be accessed at https://www.researchallofus.org/data-tools/workbench/.

### Code Availability

All code is available on GitHub to aid in reproducibility and transparency. The code can be found at https://github.com/NIH-CARD/NDD_x_meds.

## Acknowledgements

We gratefully acknowledge All of Us participants for their contributions, without whom this research would not have been possible. We also thank the National Institutes of Health’s All of Us Research Program for making available the participant data [and/or samples and/or cohort] examined in this study.

This research has been conducted using the UK Biobank Resource under application number 33601.

## Author Contributions

Analyzed data: KSL, LS, RAL, CJ, HI, MN, HL. Interpreted results: All authors.

Resource and logistics support: VE-P, KvK-L, LF, MC, AS, MN, HL. Writing and revising manuscript: All authors.

## References

1. Santiago, J. A. & Potashkin, J. A. The impact of disease comorbidities in Alzheimer’s disease. Front. Aging Neurosci. 13, 631770 (2021).

2. Santiago, J. A., Bottero, V. & Potashkin, J. A. Biological and clinical implications of comorbidities in Parkinson’s disease. Front. Aging Neurosci. 9, 394 (2017).

3. Mo, M. et al. Antidepressant use and cognitive decline in patients with dementia: a national cohort study. BMC Med. 23, 82 (2025).

4. Allott, K., Chopra, S., Rogers, J., Dauvermann, M. R. & Clark, S. R. Advancing understanding of the mechanisms of antipsychotic-associated cognitive impairment to minimise harm: a call to action. Mol. Psychiatry 29, 2571–2574 (2024).

5. Wu, C.-C., Liao, M.-H., Su, C.-H., Poly, T. N. & Lin, M.-C. Benzodiazepine use and the risk of dementia in the elderly population: An umbrella review of meta-analyses. J. Pers. Med. 13, (2023).

6. Gray, S. L. et al. Cumulative use of strong anticholinergics and incident dementia: a prospective cohort study. JAMA Intern. Med. 175, 401–407 (2015).

7. Cortes-Flores, H., Torrandell-Haro, G. & Brinton, R. D. Association between CNS-active drugs and risk of Alzheimer’s and age-related neurodegenerative diseases. Front. Psychiatry 15, 1358568 (2024).

8. Westphal Filho, F. L., et al. Statin use and dementia risk: A systematic review and updated meta-analysis. Alzheimers Dement. (N. Y.) 11, e70039 (2025).

9. Rajan, K. B. et al. Statin initiation and risk of incident Alzheimer disease and cognitive decline in genetically susceptible older adults. Neurology 102, e209168 (2024).

10. Olmastroni, E. et al. Statin use and risk of dementia or Alzheimer’s disease: a systematic review and meta-analysis of observational studies. Eur. J. Prev. Cardiol. 29, 804–814 (2022).

11. Trompet, S. et al. Pravastatin and cognitive function in the elderly. Results of the PROSPER study. J. Neurol. 257, 85–90 (2010).

12. SPRINT MIND Investigators for the SPRINT Research Group et al. Effect of intensive vs standard blood pressure control on probable dementia: A randomized clinical trial. JAMA 321, 553–561 (2019).

13. Lisibach, A. et al. Quality of anticholinergic burden scales and their impact on clinical outcomes: a systematic review. Eur J Clin Pharmacol 77, 147–162 (2021).

14. ACB Calculator. https://www.acbcalc.com/.

15. Richardson, K. et al. Anticholinergic drugs and risk of dementia: case-control study. BMJ 361, k1315 (2018).

16. Audi, S. et al. The ‘top 100’ drugs and classes in England: an updated ‘starter formulary’ for trainee prescribers. Br. J. Clin. Pharmacol. 84, 2562–2571 (2018).

17. Witowski, N. Most common beta blockers in 2023. Definitive Healthcare https://www.definitivehc.com/blog/beta-blocker-prescription-patterns (2023).

18. Vervoordt, S. M., Arnett, P., Engeland, C., Rabinowitz, A. R. & Hillary, F. G. Depression associated with APOE status and hippocampal volume but not cognitive decline in older adults aging with traumatic brain injury. Neuropsychology 35, 863–875 (2021).

19. Bartels, C. et al. Impact of SSRI Therapy on Risk of Conversion From Mild Cognitive Impairment to Alzheimer’s Dementia in Individuals With Previous Depression. Am J Psychiatry 175, 232–241 (2018).

20. Xue, L., Bocharova, M., Young, A. H. & Aarsland, D. Cognitive improvement in late-life depression treated with vortioxetine and duloxetine in an eight-week randomized controlled trial: The role of age at first onset and change in depressive symptoms. J. Affect. Disord. 361, 74–81 (2024).

21. Banerjee, S. et al. Study of mirtazapine for agitated behaviours in dementia (SYMBAD): a randomised, double-blind, placebo-controlled trial. Lancet 398, 1487–1497 (2021).

22. Tai, X. Y. et al. Association of Dementia Risk With Focal Epilepsy and Modifiable Cardiovascular Risk Factors. JAMA Neurol 80, 445–454 (2023).

23. Oh, G., Moga, D. C., Fardo, D. W. & Abner, E. L. The association of gabapentin initiation and neurocognitive changes in older adults with normal cognition. Front Pharmacol 13, 910719 (2022).

24. Association Between Gabapentin Or Pregabalin Use and Risk of Dementia - An Analysis of the National Health Insurance Research Database in Taiwan. (2022).

25. Supasitthumrong, T. et al. Gabapentin and pregabalin to treat aggressivity in dementia: a systematic review and illustrative case report: Gabapentin and pregabalin in dementia treatment. Br. J. Clin. Pharmacol. 85, 690–703 (2019).

26. Shkodina, A. D. et al. Pharmacological and non-pharmacological approaches for the management of neuropathic pain in multiple sclerosis. CNS Drugs 38, 205–224 (2024).

27. Huang, Y.-H., Pan, M.-H. & Yang, H.-I. The association between Gabapentin or Pregabalin use and the risk of dementia: an analysis of the National Health Insurance Research Database in Taiwan. Front. Pharmacol. 14, 1128601 (2023).

28. Oh, G. Y., Moga, D. C. & Abner, E. L. Gabapentin utilization among older adults with different cognitive statuses enrolled in the National Alzheimer’s Coordinating Center (2006-2019). Br. J. Clin. Pharmacol. 89, 410–415 (2023).

29. Ballard, C. et al. Quetiapine and rivastigmine and cognitive decline in Alzheimer’s disease: randomised double blind placebo controlled trial. BMJ 330, 874 (2005).

30. Cai, L. & Huang, J. Schizophrenia and risk of dementia: a meta-analysis study. Neuropsychiatr. Dis. Treat. 14, 2047–2055 (2018).

31. Shin, C. et al. Clonazepam for probable REM sleep behavior disorder in Parkinson’s disease: A randomized placebo-controlled trial. J. Neurol. Sci. 401, 81–86 (2019).

32. Gronich, N. et al. β2-adrenoceptor agonists and antagonists and risk of Parkinson’s disease: β2-ADRENOCEPTOR USE AND RISK OF PD. Mov. Disord. 33, 1465–1471 (2018).

33. Searles Nielsen, S., Gross, A., Camacho-Soto, A., Willis, A. W. & Racette, B. A. β2-adrenoreceptor medications and risk of Parkinson disease: β2-Adrenoreceptor Drugs and PD. Ann. Neurol. 84, 683–693 (2018).

34. Sittichokkananon, A., Garfield, V. & Chiesa, S. T. Genetic and lifestyle risks for coronary artery disease and long-term risk of incident dementia subtypes. Circulation 151, 1235–1247 (2025).

35. Viaggi, B. et al. Tissue penetration of antimicrobials in intensive care unit patients: A systematic review-part II. Antibiotics (Basel) 11, 1193 (2022).

36. Choi, S. et al. Association of chronic periodontitis on Alzheimer’s disease or vascular dementia. J. Am. Geriatr. Soc. 67, 1234–1239 (2019).

37. Seyedmoalemi, M. A. & Saied-Moallemi, Z. Association between periodontitis and Alzheimer’s disease: A narrative review. IBRO Neurosci. Rep. 18, 360–365 (2025).

38. World Health Organization. Pocket Book of Hospital Care for Children: Guidelines for the Management of Common Childhood Illnesses. (World Health Organization, 2013).

39. Conceição, M. et al. Repurposing doxycycline for Alzheimer’s treatment: Challenges from a nano-based drug delivery perspective. Brain Behav Immun Health 42, 100894 (2024).

40. [No title]. https://www.accessdata.fda.gov/drugsatfda_docs/label/2021/019537s092lbl.pdf.

41. [No title]. https://www.accessdata.fda.gov/drugsatfda_docs/label/2021/019537s092lbl.pdf.

42. Youssef, M. A. M., Mohamed, T. M., Bakry, A. A. & El-Keiy, M. M. Synergistic effect of spermidine and ciprofloxacin against Alzheimer’s disease in male rat via ferroptosis modulation. Int J Biol Macromol 263, 130387 (2024).

43. Salomon-Zimri, S. et al. Combination of ciprofloxacin/celecoxib as a novel therapeutic strategy for ALS. Amyotroph Lateral Scler Frontotemporal Degener 24, 263–271 (2023).

44. Shafieinouri, M. et al. Gut-brain nexus: Mapping multi-modal links to neurodegeneration at Biobank scale. medRxiv (2024) doi:10.1101/2024.09.12.24313490.

45. Schroevers, J. L. et al. Antihypertensive medication classes and risk of incident dementia in primary care patients: a longitudinal cohort study in the Netherlands. Lancet Reg. Health Eur. 42, 100927 (2024).

